# Community responses during early phase of the COVID-19 epidemic: a cross-sectional study

**DOI:** 10.1101/2020.04.04.20053546

**Authors:** Mohammad Hossein Delshad, Fatemeh Pourhaji, Fahimeh Pourhaji, Saeed Reza Ghanbarizadeh, Hassan Azhdari Zarmehri, Edris Bazrafshan, Mahdi Gholian-Aval

## Abstract

Community responses are important for outbreak management during the early phase when preventive interventions are the major options. Therefore, this study aims to examine the behavioral responses of the community during the early phase of the COVID-19 epidemic in the Razavi Khorasan Province of Iran. A cross-sectional online survey was proceeded after confirmed COVID-19 in Iran. The population of the study was 500 residents of Razavi Khorasan areas were randomly surveyed. The research tool was demographic and risk perception questionnaire and Anxiety was assessed using the 7-item GAD Scale. The data analyzed using the SPSS statistical version (V.20). The means of age participants was 31.9±11.9. The mean GAD-7 scores were 6.4±5.2 and 92.4% had moderate or severe anxiety (GAD-7 score ≥10). Many respondents reported their health status were very good or good (62.2 %; 311/500). About a quarter of them had respiratory symptoms in the past 14 days and experienced 20% of them travelled outside the Razavi Khorasan Province in the previous. Risk perception toward COVID-19 in the community of the Razavi Khorasan Province was moderate. Most participants are alert to disease progression. This study suggested timely behavioral assessment of the community is beneficial and effective to inform next intervention, and risk communication strategies in epidemic disease.

## Introduction

During the 2019–20 corona virus outbreak, Iran reported its first confirmed cases of SARS-CoV-2 infections on 19 February 2020 in Qom[1]. As of 7 March 2020, there have been 5800 confirmed cases and 145 deaths in Iran (Fig 1, 2). [2,3].

**Fig 1.**
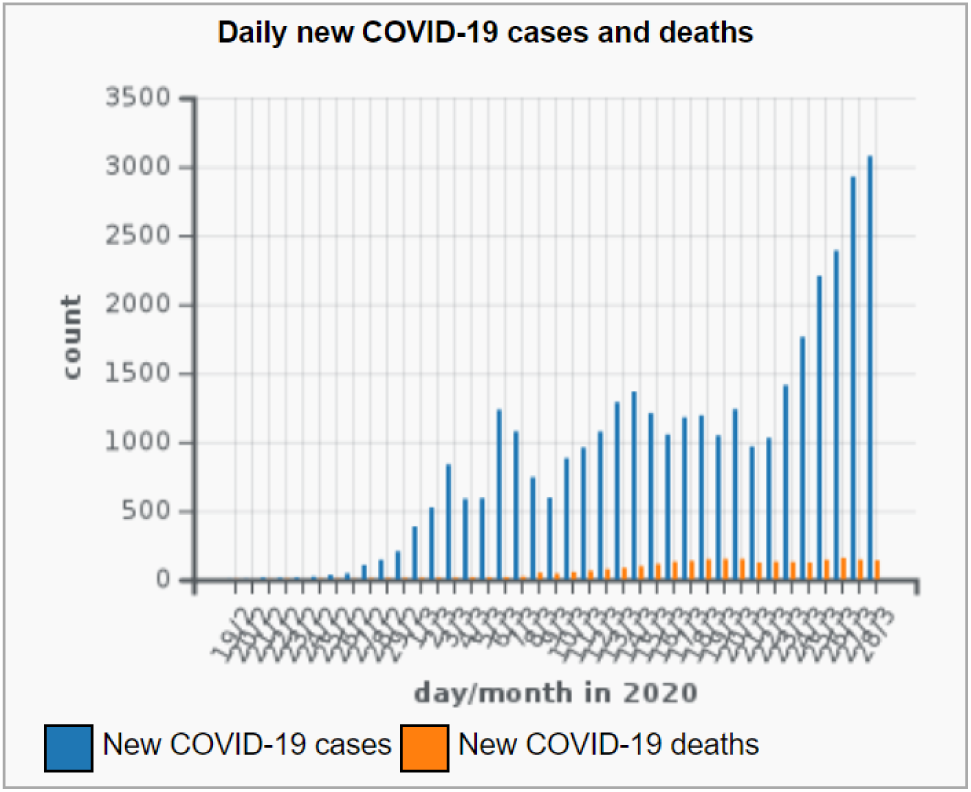
The information daily new COVID-19 cases and deaths in Iran.

**Fig 2.**
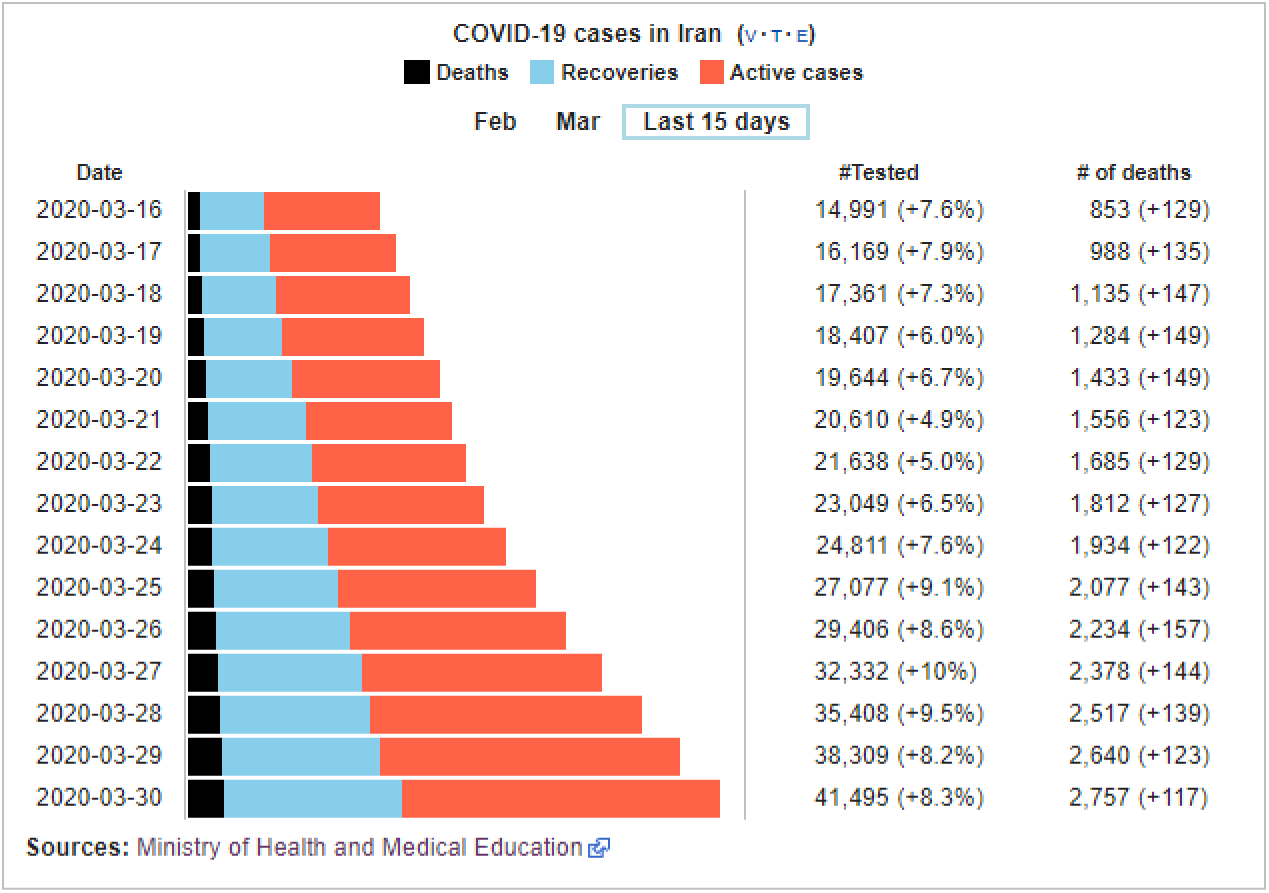
The information COVID-19 cases in Iran.

The research shows host’s behaviors are important for outbreak management, particularly during the early stage of disease and when there are not available vaccination or treatment and non-pharmaceutical intervention (NPIs)[4]. So, the efficacy of this approach depends on precautionary behaviors in people, such as hand hygiene, wearing masks and self-isolation. One of the main constructs in Health belief Model (HBM) and Protection Motivation Theory (PMT) is risk perception. Risk perception is an important factor in a voluntarily occupy in individuals. So that, people with a higher risk perception are more likely to engage in precautionary behaviors.

The previous research suggested one of the most important effects of the Corona virus outbreak is anxiety and it has led to serious concerns for citizens in all countries [5].

In addition, the growth of information technology and the acquisition of information from various sources, especially cyberspace and social networks, have made people aware of different sources and this may affect their risk perception. Given the importance of host behavior in reducing transmission and the vision timely inform in COVID-19 disease, this study aimed to investigate risk perception and behavioral responses at the early phase of the COVID-19 epidemic in Iranian population.

## Materials and Methods

A cross-sectional online survey was proceeded after confirmed COVID-19 in Iran. The population of the study was 500 residents of the Razavi Khorasan Province were randomly surveyed. The research tool was demographic (including sex, age, education and occupational status, self-perceived health status, travel history in the past month and Generalized Anxiety Disorder anxiety level. The generalized anxiety disorder questionnaire GAD [6] which measures the severity and severity of symptoms of generalized anxiety disorder over the past two weeks. Many studies have showed that GAD is valid for Iranian community population[7]. A cut point was identified that optimized sensitivity (89%) and specificity (82%)[6]. The cut- off point is scale 10.

## Risk perception

Risk perception toward COVID-19 was measured by questionnaire and two dimensions: 1) perceived susceptibility, and 2) perceived severity. Perceived susceptibility was assessment by how likely are you or your family to become infected with COVID-19 if you do not preventive behavior? The perceived severity was measured by how likely are you COVID-19 is serious, their perceived chance of having COVID-19 cured and that of survival if infected with COVID-19.

## Information sources

Participants were asked 1 question about the sources which they obtained information about COVID-19. They were included: television/radio, doctors/nurses, families or friends, newspaper, magazines, office websites, unofficial websites, social network. They were also answered 1 question about the type of information that they interested to receive.

## Preventive measures

Participants were asked whether they carry out precautionary measures: hygienic practices, Observe social distancing and travel avoidance. Participants from Razavi Khorasan Province areas were invited to share our survey link and promotion messages on their web pages, social media networks and channels usually use to transmit information for the target population and there was no limits on their emissions. Participants who were 18 or above, understood Persian and lived in Iran in the last month are eligible to participate. To avoid duplicate responses from the same responder, the survey could only be taken once from the same electronic device.

## Anxiety

Anxiety was assessed using the 7-item GAD Scale [6]. The generalized anxiety disorder questionnaire which measures the severity of symptoms of generalized anxiety disorder over the past two weeks. Many studies have showed that GAD is valid for Iranian community population[7]. A cut point was identified that optimized sensitivity (89%) and specificity (82%)[6]. The cut-off point is scale 10 and summary GAD scores ranging 0-21 [8].

## Ethical consideration

This study has been approved by the Ethics Committee of Torbat Heydariyeh University of Medical Sciences (ID:IR.THUMS.REC.1398.055).

## Statistical analysis

Frequency and proportion of participants organized. Statistical data were analyzed using independent t-test, paired t-test, one-way ANOVA and Pearson correlation coefficient and by using the SPSS statistical version (V.20).

## Results and discussion

The survey was conducted 19 Feb 2020 to 13 March 2020 (Fig 3). Our survey period covers important clinical cases, including the first local death. Data from 500 participants were analyzed. The means of age participants was 31.9±11.9. Many of the respondents were female (74.2%; 371/500). The majority were 18-27 years old (42.6%). The results indicated a 167 (33.4%) of participants were single, 327 (65.4%) married and 6 (1.2%) divorced. The mean GAD-7 scores were 6.4±5.2 and (92.4%; 462/500) had moderate or severe anxiety (GAD-7 score ≥10). Other results are in Table 1.

**Table 1.** Participant characteristics

| Characteristics | Number of respondents (%) (n=500) |
| --- | --- |
| **Sex** |  |
| Male | 129 (25.8) |
| Female | 371 (74.2) |
| **Age (years)** |  |
| 18-27 | 213 (42.6) |
| 28-37 | 144 (28.8) |
| 38-47 | 84 (16.8) |
| 48-57 | 42 (8.4) |
| 58 or above | 17 (3.4) |
| **Educational level** |  |
| Elementary and lower | 35 (7) |
| Guidance school | 18 (3.6) |
| High School and Diploma | 104 (20.8) |
| Graduate and above | 343 (68.6) |
| **Employment status** |  |
| Employee | 188 (43.6) |
| Employer | 14 (2.8) |
| Housekeeper | 218 (43.6) |
| Retired | 12 (2.4) |
| Unemployed | 68 (13.6) |
| **Living district** |  |
| Taybad | 12 (2.4) |
| Torbatjam | 29 (5.8) |
| Torbat-e Heydarieh | 113 (22.6) |
| Neyshabur | 22 (4.4) |
| Mashhad | 196 (39.2) |
| Kashmar | 28 (5.6) |
| Quchan | 20 (4) |
| Sarakhs | 24 (4.8) |
| Khaf | 24 (4.8) |
| Chenaran | 16 (3.2) |
| Gonabad | 16 (3.2) |

**Fig 3.**
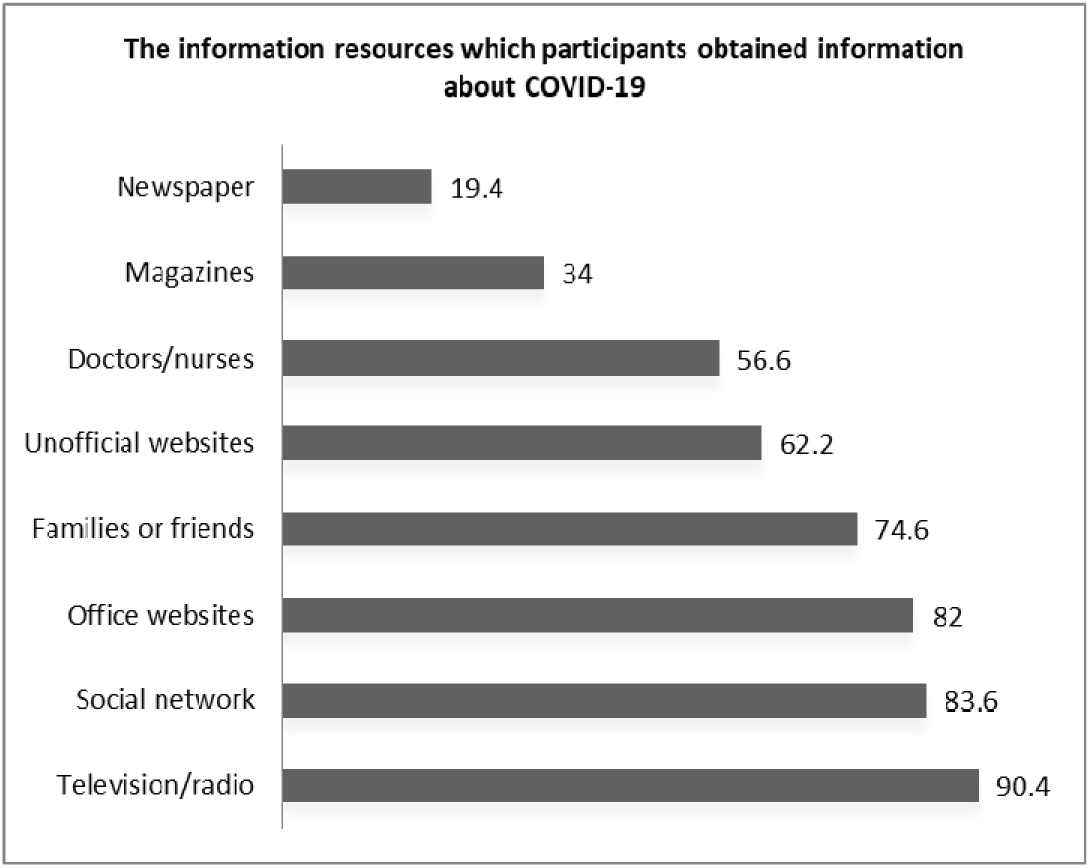
The information sources which participants obtained information about COVID-19.

Table 2 shows background in health status and travel history of participants. Many respondents reported their health status were very good or good (62.2 %; 311/500). About a quarter of them had respiratory symptoms in the past 14 days (132/500) and experienced 20% of them travelled outside Razavi Khorasan in the previous (100/500). Table 3 shows the perceived susceptibility and severity towards COVID-19 among individuals and the most participants considered themselves moderately at risk for COVID-19 (77%). The majority of respondents thought it was not likely to COVID-19. Table 4 shows the types of COVID-19 information wanted by the 500 participants who demonstrate such need. Information which participants were most interested were: number of patients infected with Corona virus (77%), Symptoms/ How to know if someone is suffering from COVID-19 (75.6%), what to do if infected with COVID-19 and the risks and consequences of COVID-19 (74%). Fig 1 shows the information sources. The most sources were television/radio (90.4%) and social network (83.6%). The results suggested the means of preventive behaviors was 20.6±4.12. Personal hygiene practices, including to avoid public transportation (61.8%), avoid eating in restaurants (64.2), avoid visiting public places (60%), wearing masks (34.8%), using gloves (37.2) and cleaning hands (54.2%). The prevalence was significantly higher in Neyshabur p≤ 0.01).

**Table 2.**
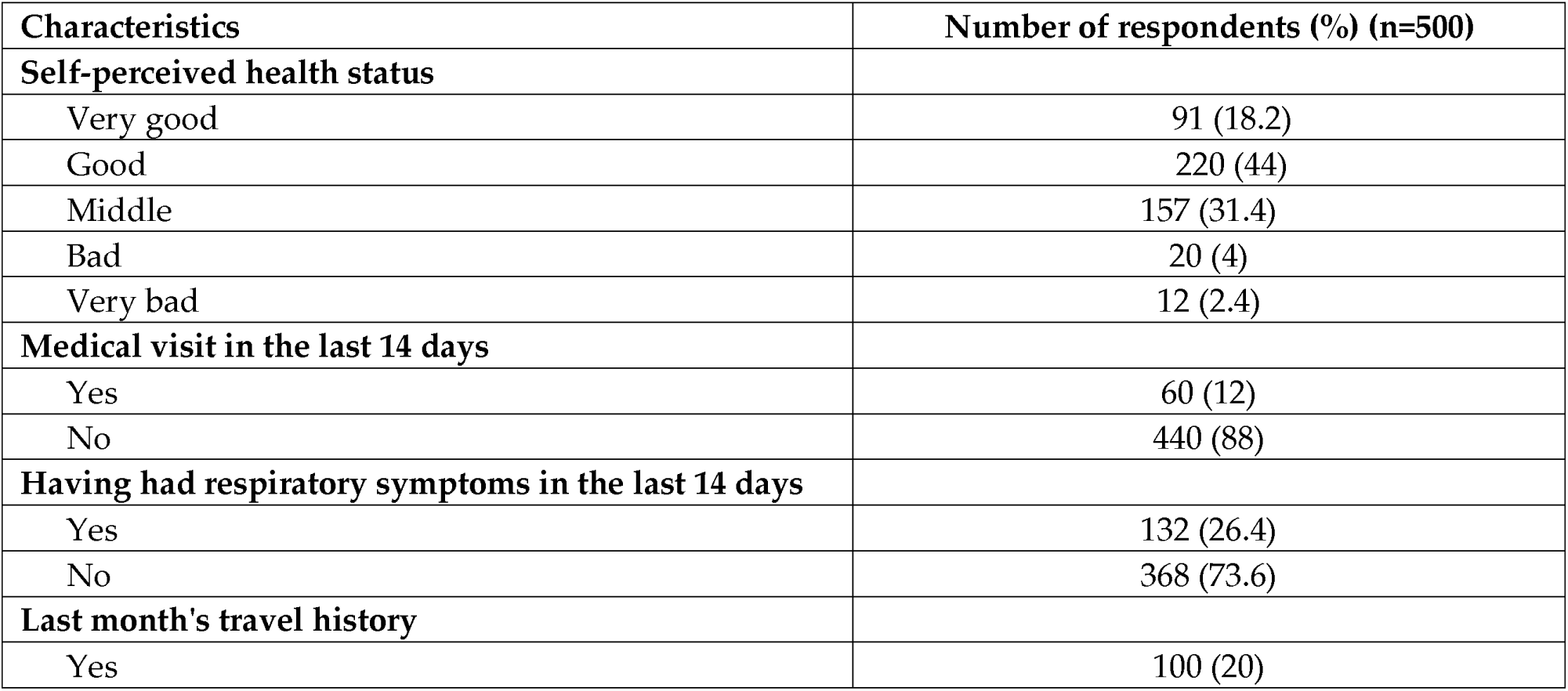
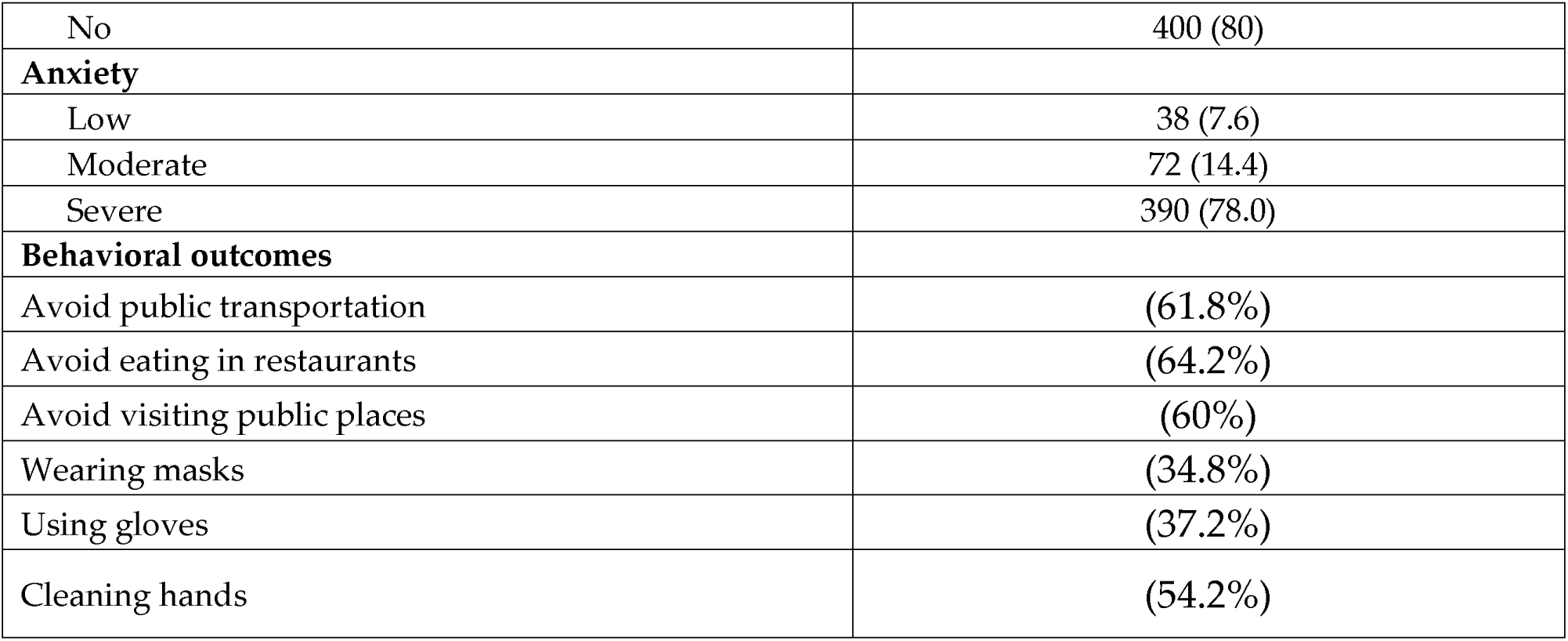
Background in health status and travel history of participants

**Table 3.**
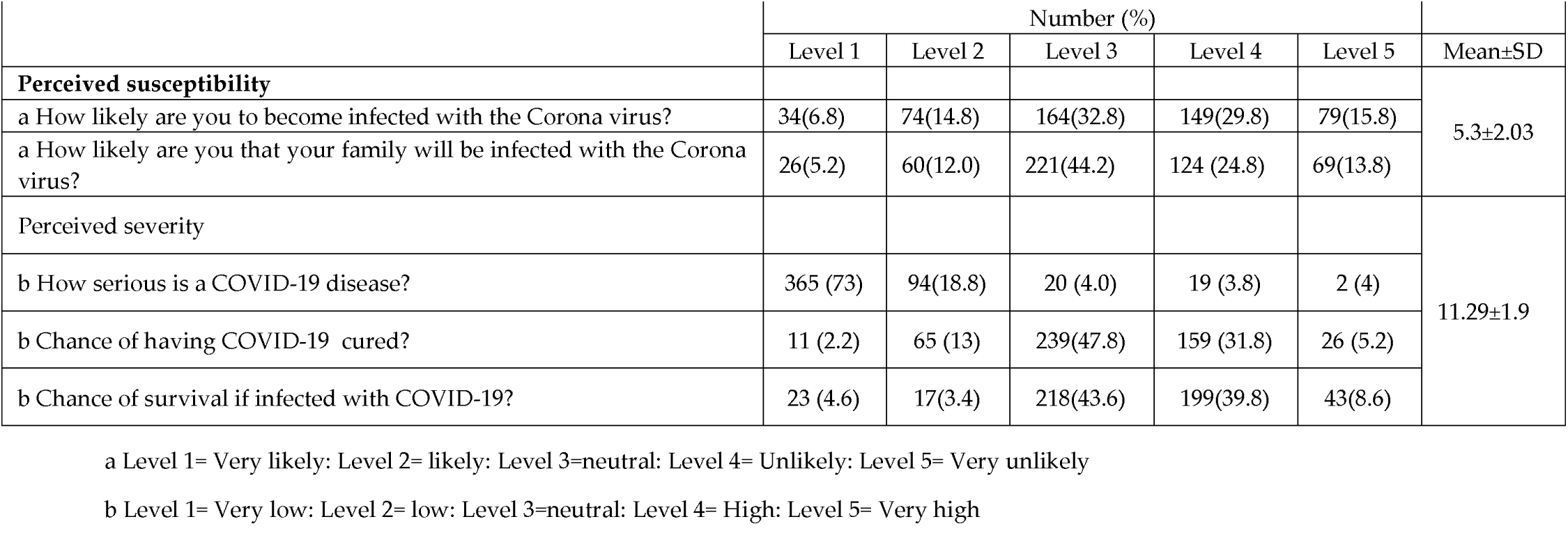
Mean and standard deviation risk perception towards COVID-19

|  | Number (%) |  |  |  |  | Mean±SD |
| --- | --- | --- | --- | --- | --- | --- |
|  | Level 1 | Level 2 | Level 3 | Level 4 | Level 5 |  |
| **Perceived susceptibility** |  |  |  |  |  | 5.3±2.03 |
| a How likely are you to become infected with the Corona virus? | 34(6.8) | 74(14.8) | 164(32.8) | 149(29.8) | 79(15.8) |  |
| a How likely are you that your family will be infected with the Corona virus? | 26(5.2) | 60(12.0) | 221(44.2) | 124 (24.8) | 69(13.8) |  |
| **Perceived severity** |  |  |  |  |  | 11.29±1.9 |
| b How serious is a COVID-19 disease? | 365 (73) | 94(18.8) | 20 (4.0) | 19 (3.8) | 2 (4) |  |
| b Chance of having COVID-19 cured? | 11 (2.2) | 65 (13) | 239(47.8) | 159 (31.8) | 26 (5.2) |  |
| b Chance of survival if infected with COVID-19? | 23 (4.6) | 17(3.4) | 218(43.6) | 199(39.8) | 43(8.6) |  |

a Level 1= Very likely: Level 2= likely: Level 3=neutral: Level 4= Unlikely: Level 5= Very unlikely
b Level 1= Very low: Level 2= low: Level 3=neutral: Level 4= High: Level 5= Very high

**Table 4.** Information required by the participants

| The information you required to receive about COVID-19 | Number (%) (n=500) |
| --- | --- |
| Distribution of COVID-19 disease cases | 290 (58) |
| Number of patients infected with Corona virus | 385 (77) |
| Interventions of Ministry of Health in Iran | 314 (62.8) |
| Preventive proceedings | 364 (72.8) |
| Disease advancement | 329 (65.8) |
| Symptoms/ How to know if someone is suffering from COVID-19 | 378 (75.6) |
| The actions of international organizations to combat COVID-19 | 312 (62.4) |
| What to do if infected with COVID-19 | 370 (74) |
| Impact of the disease in high-risk groups | 302 (60.4) |
| The Risks and Consequences of COVID-19 | 370 (74) |

This study gives a timely assessment of risk perception, information exposure, and acceptance preventive measures at an early stage of the COVID-19 epidemic in Razavi Khorasan of Iran. Although the disease was somewhat unknown to the Iranian people (including transmissibility, transmission path, and pathogenesis) at the early stage, perceived risk towards COVID-19 in the research community was moderate. In other words perceived susceptibility and perceived severity were moderate and only in 38.8% of individuals perceived susceptibility and 17.8% of the participants was high. While in the study’s Kin On Kwok and et.al [4] perceived susceptibility and severity in majority of individuals were reported high. In our study the results indicated majority of respondents thought it was not likely to COVID-19 (73%) while in china population[4] 97 percent thought it was likely to COVID-19 (97%). It seems that one of the reasons could be related to the study conditions in China that the Chinese people studied under conditions that start of large-scale social-distancing intervention, standing of sales of high-speed rail tickets to and from Wuhan, closing public places and entertainment facilities, postponing school resumption, also increasing official threat intensity in COVID-19. A great increasing generalized anxiety disorder was observed over the study period. In this study generalized anxiety disorder was 92.4%, while reported among Wuhan population 32.7% and 20.4%[8]. Also, in study of Abdulkarim Al- Rabiaah and et.al [9] generalized anxiety disorder was reported 2.7±3.1. Perhaps one reason for this is that the sample of this study was medical students who may be different in terms of information and understanding of illness and health literacy with our target group who were citizens.

Our study provides new evidence on risk perception and community responses to disease outbreaks in Razavi Khorasan to the early phase of the COVID-19 epidemic. The results suggest that the anxiety level and behaviors have changed rapidly and significantly in the early stage of the outbreak. Our finding also product several important public implications. First, providing the public with accurate and reliable information is key for addressing the psychological consequence of communicable disease outbreak. We found that the hand washing according to recommended by WHO (over 40 second) was slightly in population research. Then, further training on proper hand washing is recommended.

Our study has a number of limitations. First, in order to assess the extent to which the public responded to the major public health crisis, we shortened our survey questionnaire and shortened our survey questionnaire to obtain representative population samples using randomly sampling. Second, we asked participants to remember some of their behaviors, as a result, their answers might have recalled bias.

## Conclusions

Risk perception toward COVID-19 in the community of the Razavi Khorasan Province was moderate. Most participants are alert to disease progression. It seems that psychological and behavioral outcomes to COVID-19 had been considerable during the phase of the outbreak. The level of anxiety increased from its normal level in this area. This study suggested timely behavioral assessment of the community is beneficial and effective to inform next intervention, and risk communication strategies in epidemic disease.

## Data Availability

I share all the data in my article from manuscripts

http://delshad264.blogfa.com/

## Acknowledgement

The authors would like to thank all the individuals who took part in the study. The authors also thank Torbat Heydariyeh University of Medical Sciences for its financial support for this study.

## References

1. ”Confirmed Cases and Deaths by Country T, or Conveyance”. Worldometer. Retrieved 28 February 2020.

2. Smith JS, David (26 February 2020). “Germany ‘heading for epidemic’ as virus spreads faster outside China”. Thomson Reuters. Archived from the original on 26 February 2020. Retrieved 26 February 2020.

3. Aboulenein AFFIccafoIesTRAftooFR.

4. Kwok KO, Li KK, Chan HH, Yi YY, Tang A, et al. (2020) Community responses during the early phase of the COVID-19 epidemic in Hong Kong: risk perception, information exposure and preventive measures. https://doi.org/10.1101/2020.02.26.20028217.

5. Sadati AK, B Lankarani MH, Bagheri Lankarani K (2020) Risk Society, Global Vulnerability and Fragile Resilience; Sociological View on the Coronavirus Outbreak. Shiraz E-Med J 21: e102263. 10.5812/semj.102263.

6. Spitzer R, Kroenke K, Williams J, Bernd L (2006) A Brief Measure for Assessing Generalized Anxiety Disorder The GAD-7. Arch Intern Med: 166. 10.1001/archinte.166.10.1092.

7. Naeinian MR, Shaeiri MR, Sharif M, Hadian M (2011) To Study Reliability and Validity for A Brief Measure for Assessing Generalized Anxiety Disorder (GAD-7). Clinical Psychology & Personality 2: 41–50.

8. Qian M, Wu Q, Wu P, Hou Z, Liang Y, et al. (2020) Psychological responses, behavioral changes and public perceptions during the early phase of the COVID-19 outbreak in China: a population based cross-sectional survey. medRxiv: 2020.2002.2018.20024448. https://doi.org/10.1101/2020.02.18.20024448.

9. Al-Rabiaah A, Temsah M-H, Al-Eyadhy AA, Hasan GM, Al-Zamil F, et al. (2020) Middle East Respiratory Syndrome-Corona Virus (MERS-CoV) associated stress among medical students at a university teaching hospital in Saudi Arabia. Journal of Infection and Public Health. https://doi.org/10.1016/j.jiph.2020.01.005.

